# Dynamic and Baseline Multi-Task Learning for Predicting Substance Use Initiation in the ABCD Study

**DOI:** 10.64898/2026.04.10.26350655

**Authors:** Mengman Wei, Hanwen Zhang, Qian Peng

## Abstract

**Background:** Early initiation of substance use is linked to later adverse outcomes, and risk factors come from multiple domains and are shared across substances. In our previous work, traditional time-to-event Cox models identified individual risk factors, but these models are not designed to jointly model multiple outcomes or capture complex non-linear relationships. Multi-task learning (MTL) can leverage shared structure across related outcomes to improve prediction and distinguish common versus substance-specific predictors. However, most MTL studies rely on baseline features and focus on single outcomes, which limits their ability to capture shared risk and temporal changes. Substance use initiation is a time-dependent process that unfolds during development and reflects changing exposures over time. Baseline-only models cannot capture these changes or represent risk dynamics. Discrete-time modeling provides a practical approach by estimating interval-level initiation risk and combining it into cumulative risk at the subject level. By integrating multi-task learning with dynamic modeling, it is possible to share information across outcomes while capturing how risk evolves over time, which may improve prediction performance.

**Methods:** Using the Adolescent Brain Cognitive Development (ABCD) Study^®^ (release 5.1), we developed two complementary multi-task learning (MTL) frameworks to predict initiation of alcohol, nicotine, cannabis, and any substance use. A baseline MTL model predicted fixed-horizon (48-month) initiation using one record per participant, while a dynamic discrete-time MTL model incorporated longitudinal interval data to model time-varying risk. Both models used multi-domain environmental exposures, core covariates, and polygenic risk scores (PRS). Performance was evaluated on a held-out test set using AUROC, PR-AUC, and calibration metrics and compared with single-task logistic regression (LR). Feature importance was assessed using permutation importance and compared with Cox proportional hazards models.

**Results:** Among 2,366 unrelated participants of European genetic ancestry, initiation rates were 40.7% for alcohol, 6.4% for nicotine, 4.2% for cannabis, and 43.4% for any substance use. MTL did not consistently outperform logistic regression across all outcomes, but it showed its clearest advantages for lower-prevalence outcomes, especially cannabis and nicotine initiation. Static MTL substantially improved prediction for cannabis and nicotine compared with static logistic regression, while dynamic MTL was most useful for nicotine and cannabis in the interval-level setting. Dynamic modeling generally improved performance compared with static modeling, particularly for logistic regression and nicotine initiation, although static MTL remained stronger for cannabis. Feature-overlap analyses showed moderate agreement between static and dynamic MTL but lower concordance between MTL and Cox models. Across frameworks, behavioral and environmental predictors, especially UPPS sensation seeking and parental monitoring, were more reproducible than PRS-related features.

**Conclusions:** Dynamic multi-task learning improves the prediction of substance use initiation by leveraging longitudinal structure and shared information across outcomes. While MTL provides additional gains, incorporating time-varying information is the dominant factor for improving performance. Combining baseline and dynamic frameworks offers a comprehensive strategy for identifying robust risk factors and modeling adolescent substance use initiation.

## Introduction

Substance use often begins during adolescence, and earlier initiation is associated with increased risk of later substance use disorders and adverse psychosocial outcomes [1, 2, 3, 4]. Predicting initiation is challenging because risk arises from multiple interacting domains, including individual behavior, mental health, family environment, peers, neighborhood context, and broader sociodemographic factors [5, 6, 7]. Many of these factors are shared across substances, while some are substance-specific [8, 9, 10]. Most prior modeling approaches treat each substance outcome separately, which may fail to capture shared structure across related behaviors. Multi-task learning (MTL) provides a framework to jointly model correlated outcomes by learning shared representations while allowing task-specific prediction components [11, 12]. This approach can improve performance in settings with limited positive cases and help distinguish predictors that generalize across outcomes from those that are outcome-specific.

However, many prediction studies rely only on baseline features and do not account for how risk changes over time. Since substance use initiation unfolds longitudinally, models that incorporate temporal structure may better reflect real-world risk processes.

In this study, we propose an MTL pipeline (Figure 1) for predicting initiation of alcohol, nicotine, cannabis, and any substance use in the ABCD [13, 14]cohort using multi-domain exposures, core covariates, and polygenic risk scores (PRS) [15]. We evaluate predictive performance using discrimination and calibration metrics on a held-out test set, compare against interpretable single-task performance, and assess feature importance and cross-outcome overlap [16, 17, 18, 19, 20, 21].

**Figure 1.**
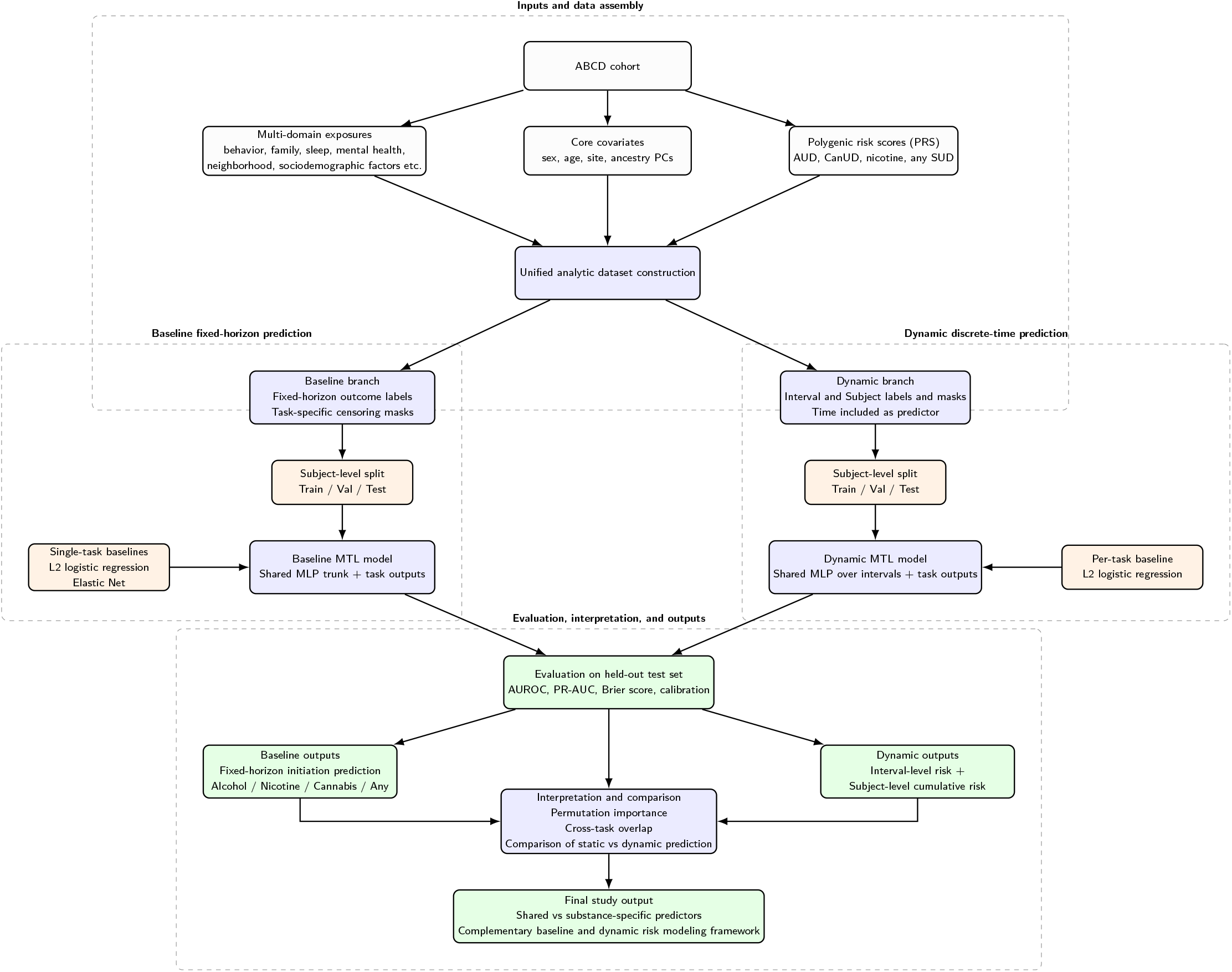
Overview of the multi-task learning pipeline for predicting substance use initiation in the ABCD Study. Multi-domain exposures, core covariates, and polygenic risk scores were assembled into a unified analytic dataset. Two complementary modeling branches were implemented: (1) a baseline fixed-horizon multi-task model using one baseline record per participant and (2) a dynamic discrete-time multi-task model using intervalized longitudinal observations. Both frameworks predicted initiation for alcohol, nicotine, cannabis, and any substance use, and were compared with logistic-regression baselines. Model performance was evaluated on held-out test data using discrimination and calibration metrics, and predictors were summarized using feature-importance and overlap analyses.s

**Figure 2.**
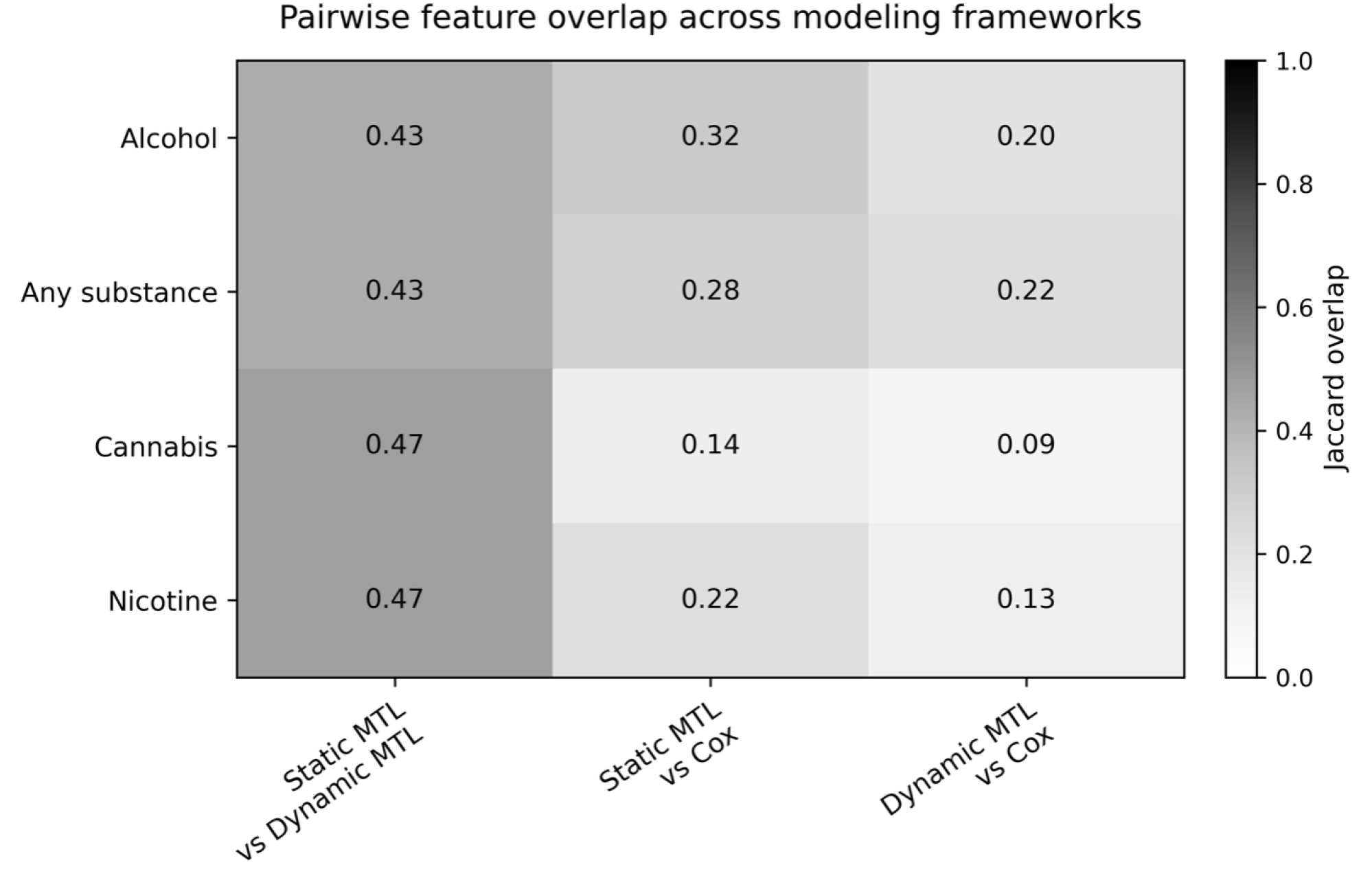
Pairwise feature overlap across static multi-task learning, dynamic multi-task learning, and Cox models. The heatmap shows the Jaccard overlap of selected or important features across modeling frameworks for alcohol, nicotine, cannabis, and any substance initiation. Static and dynamic MTL models showed moderate feature overlap across outcomes, with Jaccard indices ranging from 0.43 to 0.47. In contrast, overlap between MTL models and Cox-selected features was lower. Static MTL showed greater concordance with Cox models than dynamic MTL across all outcomes, suggesting that static baseline MTL identified feature sets more similar to those from traditional survival models, whereas dynamic MTL captured a more distinct set of longitudinal predictive signals.

**Figure 3.**
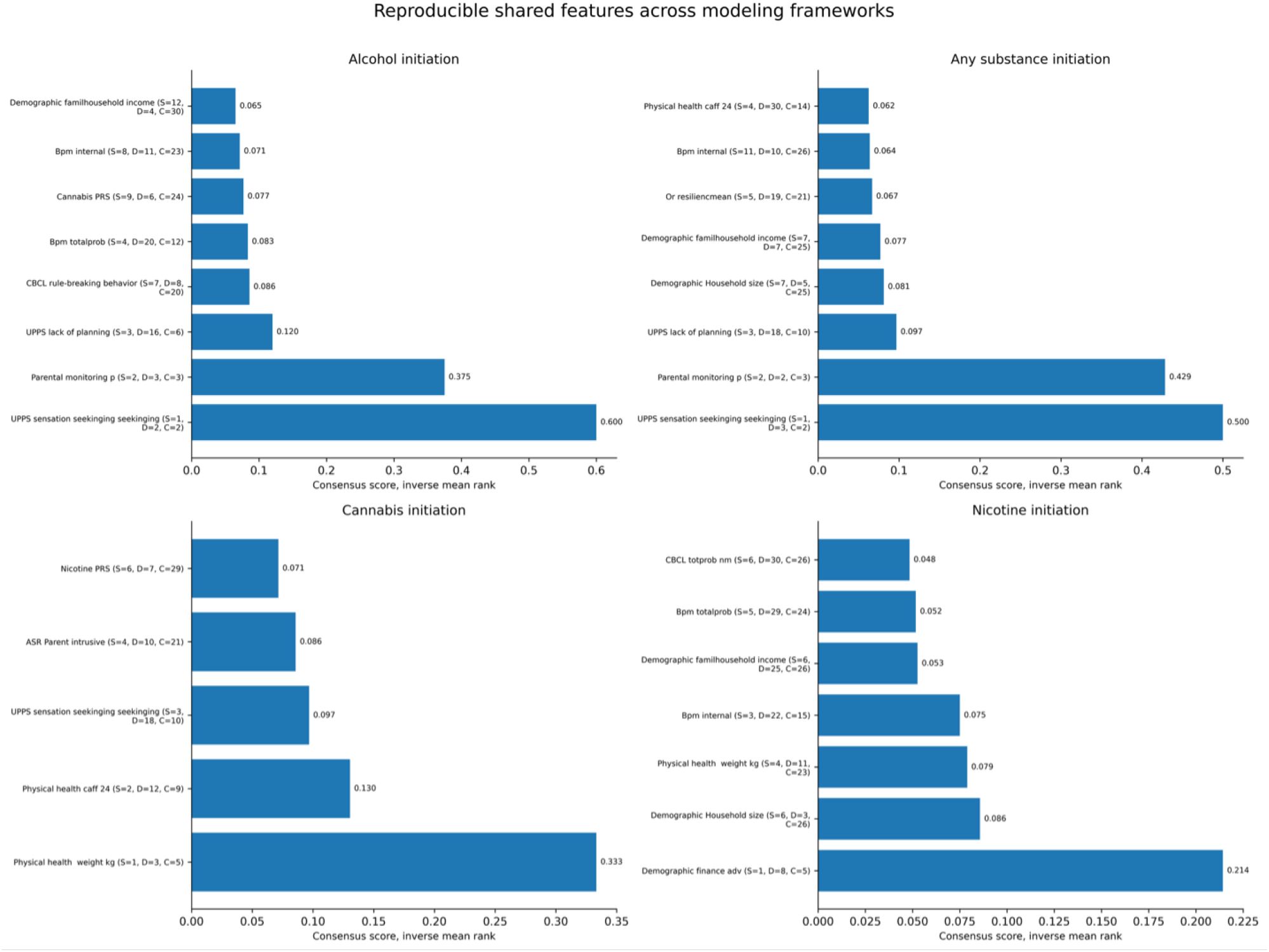
Reproducible shared features across modeling frameworks. The figure shows the top reproducible predictors for alcohol, any substance, cannabis, and nicotine initiation across modeling frameworks. Feature reproducibility was summarized using a consensus score based on inverse mean rank, for which higher values indicate stronger and more consistent ranking across models. UPPS sensation seeking and parental monitoring were among the most reproducible predictors for alcohol and any substance initiation. Cannabis and nicotine initiation showed more outcome-specific feature patterns, including physical health, demographic, and behavioral predictors. Overall, the results suggest that behavioral and environmental factors were more consistently prioritized across models than PRS-related features.

To address both shared structure and temporal dynamics, we integrate two complementary approaches: a baseline fixed-horizon MTL model and a dynamic discrete-time MTL model. The baseline model summarizes risk within a fixed prediction window, while the dynamic model represents time-varying risk across longitudinal intervals. This combined framework allows direct comparison between static and dynamic prediction strategies.

## Method

### Study Cohort

We used data from the Adolescent Brain Cognitive Development (ABCD) Study^®^ release 5.1, a large, prospective, longitudinal, multi-site study in the United States. The ABCD Study enrolled approximately 11,880 children at ages 9–10 years across 21 research sites and follows them into adolescence and early adulthood. The study includes harmonized assessments spanning neuroimaging, cognition, mental and physical health, substance use, and environmental and sociodemographic measures.

To reduce potential confounding due to population structure and relatedness, the primary analyses were restricted to unrelated participants of European genetic ancestry. Participants were included if they had valid baseline covariates, genotype-derived ancestry principal components, and longitudinal substance-use outcome data. Individuals with missing key demographic or genetic information were excluded from the final analytic cohort.

This unrelated European-ancestry cohort was used consistently across the survival, causal, and machine-learning analyses to ensure that all results were based on the same well-defined study population. Additional details are provided in our previous work [22].

### Train/validation/test splitting (subject-level)

We created a subject-level split with zero IID overlap across train/validation/test sets using a single random permutation with seed and proportions 70/15/15 (train/val/test). The split universe was defined by unique participants present in the covariate table (IID). Split manifests were saved for reproducibility.

### Baseline multi-task model

#### Baseline feature construction

We derived baseline features from the time-varying covariate table by retaining a single record per participant, defined as the earliest available visit (i.e., the minimum value of time months).

Baseline features included three components: (1) numeric exposure variables measured at baseline; (2) core covariates, including sex, age at baseline, study site, and genetic ancestry principal components (PC1–PC20); and (3) polygenic risk scores (PRS) for alcohol use disorder, cannabis use disorder, nicotine use disorder, and any substance use disorder, merged by participant identifier (IID) when available.

Missing values were handled by converting variables to numeric where possible, followed by median imputation during model training. Any remaining missing values were set to 0.

#### Outcome definition: baseline and horizon classification with censoring masks

For each substance outcome (alcohol, nicotine, cannabis, and any substance use), we used ABCD time-to-event data that include an event indicator and an event or censoring time. Because times are often recorded as absolute age in months, we converted them to follow-up time since baseline by subtracting each participant’s baseline age (in months).

We defined a binary initiation label using a fixed prediction horizon *H* months (primary *H* = 48). Participants were labeled as positive if initiation occurred before *H*. They were labeled as negative if they remained initiation-free for at least *H* months (i.e., initiation occurred after *H*, or no initiation with follow-up ≥ *H*).

If a participant was censored before *H* (no event and follow-up *< H*) or had missing event/time information, the label was treated as unobserved and excluded for that outcome. Participants were retained if they had at least one observed outcome label across all tasks.

#### Multi-task model architecture, loss, hyperparameter optimization, and early stopping

We trained a multi-task neural network with a shared multilayer perceptron (MLP) trunk and task-specific outputs for alcohol, nicotine, cannabis, and any substance use, following standard multi-task learning practice [11].

### Notation

Let *t* ∈ {1, …, *T* } index tasks and *i* ∈ {1, …, *N* } index individuals. We define:

- *m*_*it*_ ∈ {0, 1}: label-observed mask (1 if the label exists for individual *i* in task *t*),
- *α*_*t*_: task weight,
- pos weight_*t*_: positive-class weight for class imbalance,
- *z*_*it*_: model logit output,
- *y*_*it*_: observed binary label.

### Model architecture

The shared trunk used ReLU activations and dropout for regularization [23]. We evaluated two output designs: (i) a single joint output layer producing logits for all tasks, and (ii) task-specific adapter modules with separate output heads to allow limited task customization on top of shared representations [12].

### Loss function

Model training minimized masked binary cross-entropy with logits:

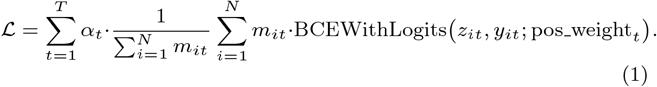

This formulation ensures that each individual contributes to the loss only for tasks with observed labels. To address class imbalance, we used task weights *α*_*t*_ and task-specific positive-class weights pos weight_*t*_ [24, 25].

### Hyperparameter optimization and training

We tuned model architecture and optimization parameters, including hidden layer sizes, dropout rate, learning rate, weight decay, batch size, and loss weights, using Optuna with the Tree-structured Parzen Estimator (TPE) sampler and optional median pruning [26].

Early stopping was applied based on a validation objective combining performance across tasks (AUROC and PR-AUC), with training stopped when improvement plateaued over a fixed patience window [27].

#### Baseline models (single-task)

As interpretable single-task baselines, we fit a separate regularized logistic regression model for each outcome (alcohol, nicotine, cannabis, and any substance use), using only participants with an observed label for the corresponding outcome.

We used L2-regularized logistic regression (ridge) to improve stability in the presence of correlated predictors [28, 29, 30].

#### Dynamic discrete-time multi-task model

##### Dynamic multi-task neural network

We trained a multi-task neural network to predict initiation risk for all outcomes at the interval level. Input features included all numeric variables, excluding identifiers and bookkeeping columns. Time variables were included as numeric predictors.

All features were coerced to numeric format. Columns that became entirely missing after coercion were removed. Missing feature values were imputed using the median, and any remaining missing values were set to 0. We standardized all features using a StandardScaler (scikit-learn) fit on the training split only, and applied the same transformation to the validation and test splits [31].

The neural network consisted of a shared multilayer perceptron (MLP) trunk with two hidden layers (256 and 128 units), ReLU activations, and dropout (rate = 0.2) for regularization [32]. A single linear output layer produced one logit per task for each interval, and predicted probabilities were obtained using the sigmoid function.

### Loss function and training

Let *z*_*it*_ denote the model logit for interval *i* and task *t, y*_*it*_ the binary label, and *m*_*it*_ ∈ {0, 1} the label mask (1 indicates an observed label). We minimized masked binary cross-entropy with logits:

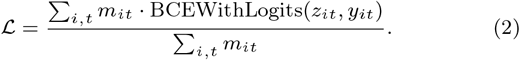

This formulation ensures that each interval contributes to the loss only for tasks with observed labels.

We used the Adam optimizer with weight decay for training. Models were trained for up to a fixed maximum number of epochs, with early stopping based on masked validation loss using a fixed patience. Gradient clipping was applied to improve training stability. The final model was selected based on the lowest validation loss and evaluated on the held-out test split.

#### Evaluation and feature importance

### Interval-level evaluation

On the test split, interval-level metrics were computed separately for each task using only intervals with observed labels (*m*_*it*_ = 1). We reported the area under the receiver operating characteristic curve (AUROC) and area under the precision–recall curve (PR-AUC). We also computed the Brier score and classification accuracy using a probability threshold of 0.5.

### Subject-level cumulative risk

To obtain subject-level predictions, we aggregated interval-level probabilities across all observed intervals for each subject in the test split (*m*_*it*_ = 1). For a subject with interval probabilities {*p*_*k*_}, cumulative risk was defined as:

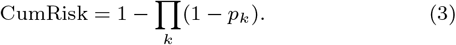

Subject-level ground truth was defined as whether the subject had at least one interval with *y*_*it*_ = 1 for the corresponding task among observed intervals. We evaluated subject-level performance using AUROC and PR-AUC.

To quantify uncertainty, we estimated 95% confidence intervals using bootstrap resampling over subjects (sampling subject IDs with replacement, repeated a fixed number of times), and computed the 2.5th and 97.5th percentiles of the bootstrap distribution.

### Baseline model (logistic regression)

As a baseline, we trained a separate L2-regularized logistic regression model for each task. For each outcome, the model was fit on training intervals with observed labels (*m*_*it*_ = 1) and evaluated on test intervals with observed labels. We reported interval-level AUROC and PR-AUC for the baseline.

### Permutation feature importance

We computed permutation feature importance on the test split as the decrease in interval-level AUROC after permuting one feature at a time. Importance was calculated separately for each task and averaged across a small number of repeats. To control computational cost, the number of evaluated features was limited to a predefined maximum.

#### Evaluation and comparison with survival and causal-style estimates

On the held-out test set, we evaluated model discrimination using the area under the receiver operating characteristic curve (AUROC) and the area under the precision–recall curve (PR-AUC), with PR-AUC emphasized due to its greater informativeness under class imbalance [33].

Calibration was assessed using the Brier score, expected calibration error (ECE), and calibration curves, to evaluate the agreement between predicted probabilities and observed event rates [34].

To characterize feature importance, we computed permutation importance as the decrease in test AUROC after randomly permuting each feature, thereby breaking its association with the outcome [35]. To place predictive importance in an inferential context, we compared top-ranked machine learning features for each outcome with results from time-varying Cox models, including hazard ratios and multiple-testing–adjusted significance, and, where available, marginal structural model (MSM) estimates. We examined the extent to which features identified as important for prediction also showed statistically supported associations in survival and causal analyses [22, 36, 37].

## Results

### Cohort Construction and Data Characteristics

For all analyses, we limited the analytic sample to unrelated participants of European genetic ancestry who had available polygenic risk scores (PRS), covariates, and longitudinal substance-use initiation data. This restriction was used to reduce possible confounding from population stratification and genetic relatedness. It also helped ensure that the target sample was consistent with the European-ancestry framework used to construct the PRS.

The final unrelated European-ancestry analytic cohort included 2,366 participants. At baseline, the mean age was 9.49 years, with a standard deviation of 0.51 years. The cohort included 1,247 male participants and 1,119 female participants, corresponding to 52.7% male and 47.3% female. We analyzed four time-to-event outcomes: alcohol initiation, nicotine initiation, cannabis initiation, and any substance initiation. During follow-up, 964 participants initiated alcohol use, 151 initiated nicotine use, 100 initiated cannabis use, and 1,027 initiated any substance use. The corresponding event rates were 40.7% for alcohol initiation, 6.4% for nicotine initiation, 4.2% for cannabis initiation, and 43.4% for any substance initiation.

Sex-specific event rates were generally similar for alcohol, cannabis, and any substance initiation. However, nicotine initiation was slightly higher among female participants. Specifically, alcohol initiation occurred in 41.6% of female participants and 39.9% of male participants. Nicotine initiation occurred in 7.6% of female participants and 5.3% of male participants. Cannabis initiation occurred in 3.9% of female participants and 4.5% of male participants. Any substance initiation occurred in 44.1% of female participants and 42.8% of male participants. Additional details are provided in our previous work [22].

#### Static and Dynamic Multi-Task Learning Versus Logistic Regression Model Performance

We first compared the performance of static multi-task learning (MTL) and logistic regression (LR) models across four substance-use initiation outcomes (Table 1). In the static setting, MTL did not uniformly outperform LR across all outcomes. For alcohol initiation, static LR showed slightly higher AUROC than static MTL (0.717 vs. 0.707), whereas static MTL showed slightly higher PR-AUC (0.614 vs. 0.606). A similar mixed pattern was observed for any substance initiation, for which LR had higher AUROC (0.711 vs. 0.693), but MTL had higher PR-AUC (0.622 vs. 0.602). In contrast, static MTL clearly outperformed static LR for the lower-prevalence cannabis and nicotine outcomes. For cannabis initiation, static MTL achieved higher AUROC (0.833 vs. 0.756) and PR-AUC (0.268 vs. 0.166). For nicotine initiation, static MTL also achieved higher AUROC (0.701 vs. 0.580) and PR-AUC (0.169 vs. 0.061). These results suggest that static MTL provided its clearest benefit for the lower-prevalence outcomes, while performance was mixed for alcohol and any substance initiation.

**Table 1.**
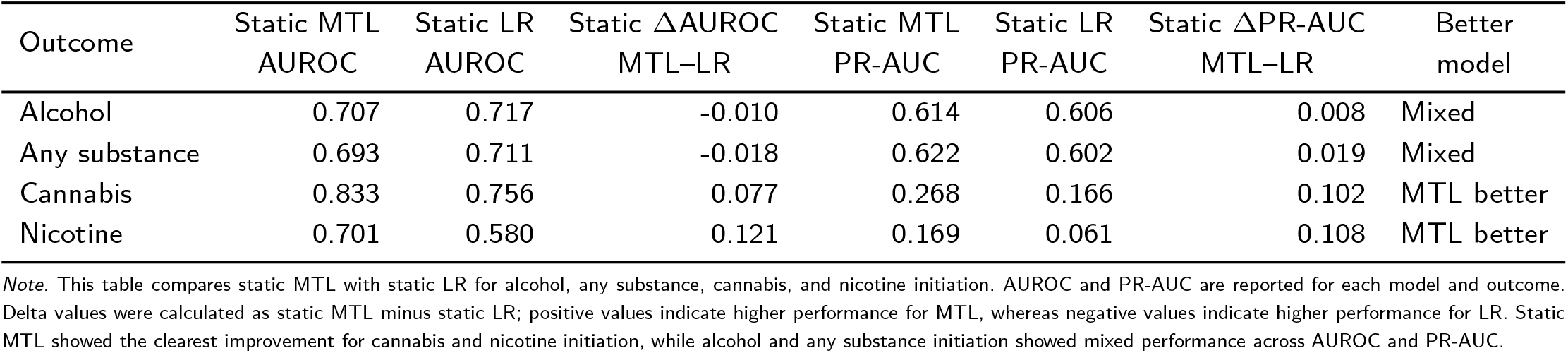
Predictive performance of static multi-task learning and logistic regression models across substance-use initiation outcomes.

| Outcome | Static MTL<br>AUROC | Static LR<br>AUROC | Static ΔAUROC<br>MTL–LR | Static MTL<br>PR-AUC | Static LR<br>PR-AUC | Static ΔPR-AUC<br>MTL–LR | Better<br>model |
| --- | --- | --- | --- | --- | --- | --- | --- |
| Alcohol | 0.707 | 0.717 | -0.010 | 0.614 | 0.606 | 0.008 | Mixed |
| Any substance | 0.693 | 0.711 | -0.018 | 0.622 | 0.602 | 0.019 | Mixed |
| Cannabis | 0.833 | 0.756 | 0.077 | 0.268 | 0.166 | 0.102 | MTL better |
| Nicotine | 0.701 | 0.580 | 0.121 | 0.169 | 0.061 | 0.108 | MTL better |

Note. This table compares static MTL with static LR for alcohol, any substance, cannabis, and nicotine initiation. AUROC and PR-AUC are reported for each model and outcome. Delta values were calculated as static MTL minus static LR; positive values indicate higher performance for MTL, whereas negative values indicate higher performance for LR. Static MTL showed the clearest improvement for cannabis and nicotine initiation, while alcohol and any substance initiation showed mixed performance across AUROC and PR-AUC.

We next evaluated dynamic MTL and dynamic LR models that incorporated longitudinal interval-level information (Table 2). In the dynamic setting, model performance again differed by outcome. For alcohol initiation, dynamic LR showed slightly higher AUROC than dynamic MTL (0.735 vs. 0.731), while dynamic MTL showed slightly higher PR-AUC (0.616 vs. 0.613). For any substance initiation, dynamic LR performed better than dynamic MTL for both AUROC (0.734 vs. 0.725) and PR-AUC (0.635 vs. 0.629). In contrast, dynamic MTL showed clearer advantages for cannabis and nicotine initiation. For cannabis initiation, dynamic MTL achieved higher AUROC (0.781 vs. 0.766), higher PR-AUC (0.196 vs. 0.188), and a slightly lower Brier score (0.035 vs. 0.036) than dynamic LR. For nicotine initiation, dynamic MTL showed a larger improvement, with higher AUROC (0.773 vs. 0.723), higher PR-AUC (0.264 vs. 0.221), and a lower Brier score (0.045 vs. 0.047). These findings indicate that dynamic MTL did not improve performance uniformly across all outcomes, but it was most useful for the rarer cannabis and nicotine initiation outcomes.

**Table 2.** Predictive performance of dynamic multi-task learning and dynamic logistic regression models across substance-use initiation outcomes.

| Outcome | Event rate | MTL mean prediction | LR mean prediction | MTL AUROC | LR AUROC | ΔAUROC MTL–LR | MTL PR-AUC | LR PR-AUC | ΔPR-AUC MTL–LR | MTL Brier | LR Brier | ΔBrier MTL–LR | Better model |
| --- | --- | --- | --- | --- | --- | --- | --- | --- | --- | --- | --- | --- | --- |
| Alcohol | 0.410 | 0.397 | 0.399 | 0.731 | 0.735 | -0.003 | 0.616 | 0.613 | 0.003 | 0.201 | 0.199 | 0.002 | Mixed |
| Any substance | 0.432 | 0.418 | 0.417 | 0.725 | 0.734 | -0.009 | 0.629 | 0.635 | -0.007 | 0.206 | 0.203 | 0.003 | Mixed |
| Cannabis | 0.041 | 0.025 | 0.030 | 0.781 | 0.766 | 0.015 | 0.196 | 0.188 | 0.009 | 0.035 | 0.036 | -0.001 | MTL useful |
| Nicotine | 0.054 | 0.057 | 0.060 | 0.773 | 0.723 | 0.049 | 0.264 | 0.221 | 0.043 | 0.045 | 0.047 | -0.002 | MTL useful |

*Note.* This table compares dynamic MTL with dynamic LR using interval-level longitudinal prediction. Event rates and mean predicted probabilities are included to assess calibration. Brier scores are reported as prediction-error metrics, for which lower values indicate better performance. Delta values were calculated as dynamic MTL minus dynamic LR. Positive delta values for AUROC and PR-AUC indicate better performance for dynamic MTL, whereas negative delta values for the Brier score indicate lower prediction error for dynamic MTL. Dynamic MTL showed the strongest advantage for cannabis and nicotine initiation, while performance was mixed for alcohol and any substance initiation.

To assess the contribution of longitudinal information, we then compared dynamic versus static models within each modeling framework, without directly comparing MTL against LR (Table 3). For MTL, dynamic modeling improved AUROC and PR-AUC for alcohol, any substance, and nicotine initiation. The largest MTL gain was observed for nicotine, for which AUROC increased from 0.701 to 0.773 and PR-AUC increased from 0.169 to 0.264. However, cannabis was an exception: static MTL outperformed dynamic MTL for cannabis initiation, with higher AUROC (0.833 vs. 0.781) and PR-AUC (0.268 vs. 0.196). For LR, dynamic modeling improved both AUROC and PR-AUC across all four outcomes. The largest LR gains were also observed for nicotine, for which AUROC increased from 0.580 to 0.723 and PR-AUC increased from 0.061 to 0.221. These results suggest that longitudinal modeling generally improved prediction, especially for nicotine, but the benefit was outcome-specific and not universal across all model families.

**Table 3.** Comparison of dynamic versus static prediction performance within multi-task learning and logistic regression models.

| Outcome | Static MTL AUROC | Dynamic MTL AUROC | $\Delta$ MTL AUROC dynamic–static | Static MTL PR-AUC | Dynamic MTL PR-AUC | $\Delta$ MTL PR-AUC dynamic–static | Static LR AUROC | Dynamic LR AUROC | $\Delta$ LR AUROC dynamic–static | Static LR PR-AUC | Dynamic LR PR-AUC | $\Delta$ LR PR-AUC dynamic–static | Interpretation |
| --- | --- | --- | --- | --- | --- | --- | --- | --- | --- | --- | --- | --- | --- |
| Alcohol | 0.707 | 0.731 | 0.025 | 0.614 | 0.616 | 0.002 | 0.717 | 0.735 | 0.017 | 0.606 | 0.613 | 0.007 | Dynamic higher for all metrics |
| Any substance | 0.693 | 0.725 | 0.032 | 0.622 | 0.629 | 0.007 | 0.711 | 0.734 | 0.023 | 0.602 | 0.635 | 0.033 | Dynamic higher for all metrics |
| Cannabis | 0.833 | 0.781 | -0.051 | 0.268 | 0.196 | -0.071 | 0.756 | 0.766 | 0.010 | 0.166 | 0.188 | 0.022 | Static MTL; dynamic LR higher |
| Nicotine | 0.701 | 0.773 | 0.071 | 0.169 | 0.264 | 0.095 | 0.580 | 0.723 | 0.143 | 0.061 | 0.221 | 0.160 | Dynamic higher for all metrics |

*Note.* This table evaluates whether adding longitudinal information improved prediction within each model family. Comparisons are made separately for MTL and LR; no direct MTL-versus-LR comparison is made. Delta values were calculated as dynamic minus static performance within the same model family. Positive values indicate higher performance for the dynamic model, whereas negative values indicate higher performance for the static model. Dynamic modeling improved LR performance across all four outcomes and improved MTL performance for alcohol, any substance, and nicotine initiation. Cannabis was the exception for MTL, for which static MTL had higher AUROC and PR-AUC than dynamic MTL.

Taken together, these analyses support three main findings First, MTL provided the clearest advantage for the lower-prevalence cannabis and nicotine outcomes, particularly in the static setting and for dynamic nicotine prediction. Second, dynamic modeling generally improved prediction compared with static modeling, especially for LR and for nicotine initiation. Third, the benefit of dynamic MTL was outcome-specific: dynamic MTL improved over static MTL for alcohol, any substance, and nicotine initiation, but static MTL remained stronger for cannabis. Therefore, the main conclusion is not that one modeling strategy dominated across all outcomes, but that different modeling approaches captured different aspects of substance-use initiation risk. Dynamic MTL appears most useful for selected lower-prevalence outcomes, whereas dynamic LR remained competitive or stronger for broader outcomes such as alcohol and any substance initiation.

#### Feature Importance Across MTL Models and Concordance With Traditional Cox Models

To evaluate whether machine-learning models identified predictors consistent with traditional survival models, we compared the overlap of important features across static MTL, dynamic MTL, and Cox-based models. Feature overlap was quantified using the Jaccard index separately for alcohol, nicotine, cannabis, and any substance initiation.

Overall, the static and dynamic MTL models showed moderate overlap in selected features across all four outcomes. The Jaccard overlap between static MTL and dynamic MTL ranged from 0.43 to 0.47, with the highest overlap observed for cannabis and nicotine initiation. This suggests that both MTL approaches captured a partially shared set of predictive features, although each model also selected outcome-relevant features that were not identified by the other approach.

In contrast, concordance between the MTL models and Cox models was lower. Static MTL showed modest overlap with Cox-selected features, with Jaccard indices of 0.32 for alcohol, 0.28 for any substance, 0.14 for cannabis, and 0.22 for nicotine. Dynamic MTL showed even lower overlap with Cox models, with Jaccard indices of 0.20 for alcohol, 0.22 for any substance, 0.09 for cannabis, and 0.13 for nicotine.

Across outcomes, static MTL consistently showed greater feature overlap with Cox models than dynamic MTL. This pattern indicates that static baseline MTL may recover feature sets more similar to those identified by traditional survival modeling, whereas dynamic MTL may prioritize features that improve longitudinal prediction but are less directly aligned with Cox-based associations. The weakest concordance was observed for cannabis initiation, especially between dynamic MTL and Cox models, suggesting that cannabis-related prediction may be more model-dependent or influenced by features not strongly captured by traditional Cox selection.

Together, these findings suggest that MTL models and Cox models provide complementary perspectives. Cox models identify variables with interpretable time-to-event associations, while MTL models identify features that contribute to multi-outcome prediction. The moderate overlap between static and dynamic MTL, combined with their limited overlap with Cox models, highlights the importance of integrating traditional statistical models and machine-learning approaches when studying predictors of adolescent substance-use initiation.

#### Reproducible Shared Features Across Modeling Frameworks

To identify predictors that were consistently prioritized across modeling frameworks, we summarized reproducible shared features using a consensus score based on inverse mean rank. This approach highlights features that were repeatedly ranked as important across models rather than features that appeared in only one framework.

Across outcomes, several behavioral and environmental predictors showed reproducible importance. For alcohol initiation, the strongest shared predictors included UPPS sensation seeking, parental monitoring, and UPPS lack of planning. UPPS sensation seeking had the highest consensus score for alcohol initiation, suggesting that impulsivity-related traits were among the most reproducible predictors across modeling approaches.

For any substance initiation, a similar pattern was observed. UPPS sensation seeking and parental monitoring were again the most strongly reproducible features, followed by additional behavioral, demographic, and physical health-related variables. This suggests that general substance-initiation risk may be influenced by a combination of sensation seeking, family monitoring, and broader contextual factors.

For cannabis initiation, the reproducible feature set was somewhat different. Physical health-related variables and UPPS sensation seeking appeared among the top shared predictors. The strongest consensus features showed lower scores than those observed for alcohol and any substance initiation, suggesting weaker cross-model reproducibility for cannabis-related predictors.

For nicotine initiation, reproducible features included demographic and contextual variables, physical health-related measures, and behavioral traits. Compared with alcohol and any substance initiation, nicotine showed a more distributed feature-importance pattern, with no single predictor dominating as strongly as UPPS sensation seeking did for alcohol and any substance initiation.

Overall, the reproducible shared-feature analysis suggests that behavioral traits, especially sensation seeking and related impulsivity measures, together with family and environmental context, were more consistently identified than PRS-related features. These findings support the broader result that adolescent substance-use initiation was more strongly characterized by behavioral and environmental predictors than by genetic risk scores alone.

## Discussion

This study integrates baseline and dynamic multi-task learning (MTL) approaches to model substance-use initiation in adolescence. These two frameworks provide complementary views of risk. Baseline models summarize risk within a fixed time window, while dynamic models capture how risk changes over time.

Our results show that the benefit of MTL depends on both outcome prevalence and modeling setting. In the baseline setting, MTL provided modest improvements over logistic regression (LR), with larger gains for low-prevalence outcomes such as cannabis and nicotine initiation. This is consistent with the ability of MTL to share information across related tasks when data are limited. For more common outcomes, such as alcohol and any substance initiation, performance differences between MTL and LR were small, suggesting that well-regularized single-task models already capture much of the available signal.

Incorporating longitudinal information generally improved predictive performance, although the magnitude and direction of improvement depended on both the outcome and model family. Dynamic modeling improved AUROC for LR across all four outcomes and for MTL across alcohol, any substance, and nicotine initiation; cannabis was the exception, for which static MTL remained stronger. The largest improvements were observed for nicotine initiation. These findings indicate that temporal information is a major driver of performance improvement. Modeling risk as a time-dependent process allows the model to capture evolving exposures and better reflect the underlying developmental process of substance initiation.

The dynamic MTL framework also provides practical advantages. By modeling interval-level risk and aggregating it into subject-level cumulative risk, it produces predictions that are more closely aligned with real-world risk trajectories. This formulation permits direct interpretation of how risk accumulates over time, which is not possible in baseline-only models.

Feature-importance analyses further highlight differences between modeling approaches. Agreement across methods was modest, reflecting differences in model objectives. MTL focuses on predictive performance, while Cox models emphasize statistical association. Static MTL showed greater agreement with Cox-selected features than dynamic MTL, suggesting that static baseline MTL recovered feature sets more similar to those identified by traditional survival analysis, whereas dynamic MTL captured a more distinct set of longitudinal predictive signals. Across modeling frameworks, behavioral traits, parental monitoring, physical health-related measures, and developmental or contextual factors were repeatedly prioritized, indicating a partially stable core risk structure.

These findings suggest that MTL should not be viewed as universally superior to simpler models. Instead, its benefits depend on the data structure and prediction goal. When outcomes are imbalanced or related, MTL may provide advantages. However, incorporating longitudinal information may be more important than model complexity alone.

Overall, this study supports the use of dynamic multi-task learning as a practical extension of traditional models for predicting substance-use initiation, particularly when integrating multi-domain and time-varying risk factors.

## Conclusion

This study presents a unified framework combining baseline and dynamic multi-task learning to predict substance-use initiation. The results show that incorporating longitudinal information generally improves predictive performance, with the largest gains observed for nicotine initiation, while multi-task learning provides additional benefits particularly for lower-prevalence outcomes. However, these gains were outcome-specific, and static MTL remained stronger for cannabis initiation.

Dynamic modeling captures how risk evolves over time and produces subject-level cumulative-risk estimates that are more closely aligned with real-world processes. At the same time, baseline models provide a simpler summary of risk within a fixed horizon. Together, these approaches offer complementary insights.

Across modeling frameworks, a set of consistent predictors— including behavioral traits, family environment, and developmental or contextual factors—emerged as robust risk factors. Most other features were model-specific, highlighting the importance of using multiple approaches to identify stable signals.

Overall, combining dynamic modeling with multi-task learning provides a flexible and effective strategy for predicting substance-use initiation and identifying key risk factors in longitudinal data.

## Data Availability

This study uses data from the Adolescent Brain
Cognitive Development (ABCD) Study (https://abcdstudy.org),
available through the NIMH Data Archive (NDA). The ABCD
data release used was version 5.1. The study is supported by
the National Institutes of Health (NIH) and additional federal partners under multiple award numbers, including U01DA041048
and U01DA050987. A full list of funders is available at https://abcdstudy.org/federal-partners.html.

## Data availability

### Code

The analysis code and scripts used in this study are publicly available at: https://github.com/mw742/ABCD-MultitasksML and https://github.com/mw742/ABCD-MultitasksML-dynamic.

### Data

This study uses data from the Adolescent Brain Cognitive Development (ABCD) Study (https://abcdstudy.org), available through the NIMH Data Archive (NDA). The ABCD data release used was version 5.1. The study is supported by the National Institutes of Health (NIH) and additional federal partners under multiple award numbers, including U01DA041048 and U01DA050987. A full list of funders is available at https://abcdstudy.org/federal-partners.html.

## Author contributions statement

Mengman Wei conceived the study, designed the analytical framework, performed data processing, statistical analyses, and computational modeling, and drafted the manuscript. Mengman Wei also carried out code implementation, data curation, and interpretation of the results. Hanwen Zhang contributed to data curation, visualization, and manuscript discussion. Qian Peng provided supervision, overall guidance, resource support, and funding acquisition. All authors contributed to the discussion and revision of the manuscript.

## Preprint Notice

This manuscript is a preprint and has not yet undergone peer review. The content is shared to disseminate findings and establish a precedent. Additional analyses and revisions may be incorporated in future versions.

